# Situational judgment test in A Sample of Iraqi Doctors

**DOI:** 10.1101/2023.12.22.23300482

**Authors:** Dounia Ahmed Raoof, Hawraa Haitham Kadhim, Dalal Ali Fadhil, Ali Mohammed Jawad Al-mothaffar

## Abstract

**Study Rationale:** Health Authorities expect physicians to behave professionally with serious lapses in conduct that adversely impact the patients, the public and the profession.

**Objectives:** To explore the behavior and attitude towards complex, real-life situations met in the workplace in a sample of Iraqi doctors .The question was, is there any difference in the score between rotator doctors and senior resident doctors?

**Methods:** A cross sectional study conducted for rotators and senior residents selected randomly and the questionnaire was via face to face interview.. The questionnaire contains 10 Case Scenarios taken from the UK foundation program, medical school council 2012. Each question has eight choices, and only three choices must be answered. Each correct choice is given 4 marks and the maximum mark for each question is 12, the wrong answer does not cancel the correct answer. Analysis was done using SPSS 25.0

**Results:** A sample of 125 individuals participated in this study in which 51% are males while 49% are females, and 48% of them are rotators and 52% are senior residents. The priority of patient care had the highest mean score of 7.26 out of 12 while “Communication skills” had a lower mean score of 3.74. Females tended to score higher in “Conflicts between professional and personal concerns” than males, with a mean score of 4.98 compared to 3.69.Rotators have a mean score of 57.6 out of 120 compared to 53.6 for senior residents (p = 0.048).

**Conclusion:** This study tried to explore behavior and attitude toward complex and real time situations in work place. Mean score of conflicts between professional and personal concerns was higher for females. On the other hand regarding training level, there were two items significantly different, namely conflicts between personal and professional concerns and ensuring patient safety. Communication skills had low score for both genders and training level.

## Introduction

During World War II, military selection psychologists were in need of a tool to select competent soldiers to join the armed forces. They developed a job test that consisted of detailed and realistic descriptions of challenging military situations. All descriptions were situations that armed forces were likely to encounter while on the job. After reading each situation, recruits were presented with several potential reactions to the given threat or challenge and they were asked which reaction they considered the most effective response (1). The instrument turned out to be a success. This instrument can be considered one of the first situational judgment tests (SJTs). After World War II, several similar tests were designed to capture supervisory potential (2)(3)(4). In 1990, Motowidlo and colleagues framed the SJT as a new alternative measurement procedure for personnel selection (5) and thereby reinvigorated interest in SJTs among scientists and practitioners. SJTs present test-takers with realistic job situations, followed by potential response options out of which candidates have to select the most appropriate response (5). SJTs are considered measurement tools that aim to capture job-related competencies and skills (6).

Traditionally, students and trainees within medicine have tended to be assessed on academic ability alone. This approach however has a number of limitations, and recently there has been an increasing emphasis in medical education and training on assessing for non-academic and professional attributes that are important for competent performance in clinical practice (7). In the UK for example, there are practical limitations of selecting on the basis of academic ability alone.

Although academic attainment has been shown to be a good predictor of early performance in medical school (8), longitudinal research shows that the predictive power of academic attainment declines as trainees move into clinical practice and beyond (8)(9)(10). These findings emphasize that academic ability is a necessary, but not sufficient condition to ensure that trainees become competent health professionals, and thus the importance of selecting trainees on a range of non-academic attributes in addition.

Within UK, although the values and behaviors expected of health and social care professionals are preserved in the National Health Service (NHS) Constitution (2013), recent government enquiries (11) have highlighted major concerns about the decline in compassionate care within all healthcare roles, which has global significance. These enquiries, although UK-based, have relevance internationally, as they highlight the critical role that the workforce plays in ensuring the provision of high quality and safe healthcare services and, in particular, the impact of staff values and behaviors on the quality of patient care and thus patient outcomes Undoubtedly, an important first step is ensuring that the right individuals with the appropriate values to work in clinical practice are appointed to any educational course, training place or healthcare role.

Analysis research provides supporting evidence for the importance of non-academic attributes for successful performance in various healthcare roles. For example, attributes such as empathy, resilience, team involvement and integrity are necessary for medicine and dental students (12) and in postgraduate medical training (13) (14).

In UK, SJTs are used for postgraduate training selection for a variety of roles including Public Health (15), Psychiatry (16) and Ophthalmology(16). SJTs are also used for a variety of other healthcare professions including dental foundation training (12) and veterinary science (17).

The use of SJTs in healthcare selection is expanding globally. Internationally, SJTs have been used in medical school admissions in Belgium (18), Singapore (19), Canada (20) and in postgraduate recruitment in Australia to select trainees for entry to training in general practice (21) (22).

In general, the literature has supported the criterion-related validity of SJTs. For instance, in a meta-analysis, found SJTs to have an average observed validity of 0.20 for predicting job performance. Furthermore, research has demonstrated that SJTs show smaller ethnic score differences than cognitive measures (23) and have high face and content validity (6), making SJTs an attractive selection tool.Despite their popularity and criterion-related validity, SJTs have a clear limitation: they are easy to fake by candidates in high-stakes selection settings. For instance, it has been conducted a between-subjects study on the fakability (18) of SJTs and found that candidates in the faking condition scored 0.89 SD higher than candidates in the honest condition. Faking on a selection test can be defined as a candidate’s conscious distortion of their answers to score more favorably.

A seemingly easy way to prevent faking on SJTs is to change the response instruction. There are two common types of response instructions: should-do and would-do (i.e., behavioral tendency) instructions (24). Should-do response instructions ask the candidate to identify the best or correct course of action. Would-do response instructions ask the candidate to indicate how he or she would likely behave (25). A study found that candidates can easily distort their answers on a would -do SJT (26). However, the results for the should-do SJT were inconsistent, due to the difficulty to fake knowledge. Faking even led to lower scores when candidates first answered honestly because they had already responded to the best of their ability the first time they were presented with the job dilemmas.

However, changing the response instruction to a should-do SJT is not possible without changing the constructs that are being measured with the SJT. Indeed, a study showed that construct validity is “dramatically affected by the type of instructions” (27). In general, should-do SJTs tap more into ability and knowledge related constructs and would-do SJTs tap more into attitudes and personality related constructs (25).

The aim was to to explore the behavior and attitude towards complex, real-life situations met in the workplace in a sample of Iraqi doctors .The question was, is there any difference in the score between rotator doctors and senior resident doctors?

## Subjects and Methods

This cross-sectional study was conducted in Baghdad Teaching Hospital,Al-Kadhimiya Teaching Hospital, Al-Kindi Teaching Hospital, Sheikh Zayed Hospital, Al-Yarmouk Teaching Hospital, Child welfare Hospital, Ibn Al-Nafis Hospital, during the period from June 2023 to December 2023

The study recruited senior residents and newly graduated rotators. A convenient sample of target doctors who are available at the time of data collection and agreed to participate.

### Data Collection Form and Procedure

Self-administered test form distributed to all participants consisting of 10 questions test different disciplines include “Conflicts between professional and personal concerns, Prescription error, Making decisions in a stressful situations, the priority is the patient care, communication skills, teamwork, ensuring patient safety, work under pressure and ensure the safety of yourself and others, Respecting patient confidentiality, Patient safety “. The questions were taken from the UK foundation program, medical school council 2012, each question has eight choices, and only three choices must be answered. Each correct choice is given 4 marks and the maximum mark for each question is 12, the wrong answer does not cancel the correct answer. Participants were given 20 minutes to answer the 10 questions, the questionnaire was neglected if participant answered in less than 10 minutes, then the answers collected and scored.

A pilot study of 10 doctors was done, participation rate was 100% and few modification for the set questions were done.

### Data Processing and Analysis

Data processing, Tables, graphs & calculation was done using IBM SPSS 25 package .Differences with a p-value <0.05 will be considered as statistically significant. Statistical tests were used accordingly including Chi-square and student-t tests. 157 samples were taken 32 of them have been neglected For reasons including not answering all the questions, or answering with two or four choices instead of three.

### Ethical Approval

Ethical approval was obtained from the Scientific Committee/College of Medicine/University of-Baghdad.

### Human subject protection

Before asking of the participant’s verbal consent, they were given brief description about the objectives of the study, and how to answer the questions correctly. The confidentiality of the name was observed and identification numbers were used instead of it.

## Results

In this study, the gender and grade of training distribution (Table 1) revealed a relatively balanced representation of male and female participants, with 48% of the participants identified as rotators and 52% as senior residents.

**Table 1:**
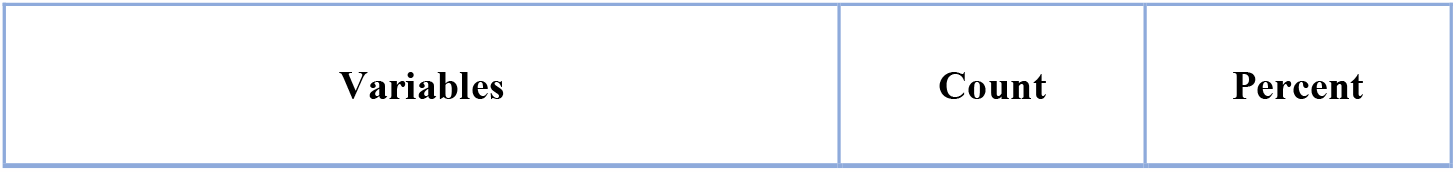
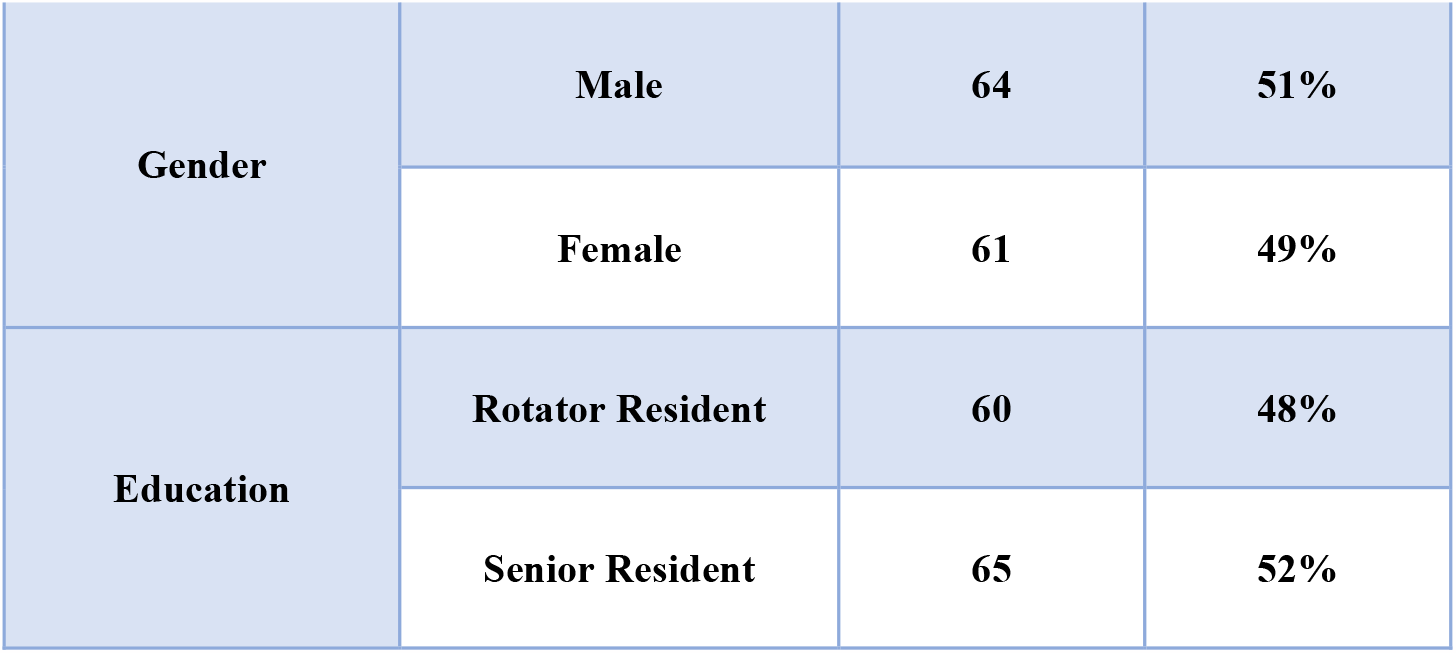
Gender and grade of training distribution.

| Variables | Count | Percent |
| --- | --- | --- |

|  |  |  |  |
| --- | --- | --- | --- |
| **Gender** | **Male** | **64** | **51%** |
|  | **Female** | **61** | **49%** |
| **Education** | **Rotator Resident** | **60** | **48%** |
|  | **Senior Resident** | **65** | **52%** |

The gender distribution within these 2 groups (Table 2) highlighted that 30% of female participants were rotators, whereas 34% of male participants were senior residents. There was a higher proportion of senior residents in the study.

**Table 2:** Distribution of gender among grade of training of participants.

| Gender | Education |  |  |  | Total |
| --- | --- | --- | --- | --- | --- |
|  | Rotator Resident |  | Senior Resident |  |  |
| Male | 22 | 18% | 42 | 34% | 64 |
| Female | 38 | 30% | 23 | 18% | 61 |
| Total | 60 | 48% | 65 | 52% | 125 |

Table 3 provides insights into the mean score distribution according to various items, Notably, “The priority is patient care” had the highest mean score of 7.26.On the other hand, “Communication skills” had a lower mean score of 3.74

**Table 3:** Mean score distribution according to items.

| **Descriptive Statistics** | **Disciplines** | **Minimum** | **Maximum** | **Mean** | **Std. Deviation** |
| --- | --- | --- | --- | --- | --- |
| **Q1- Score** | **Conflicts between professional and personal concerns** | **0** | **12** | **4.32** | **+/-2.669** |
| **Q2- Score** | **Prescription error** | **0** | **12** | **5.25** | **+/-2.755** |
| **Q3- Score** | **Making decisions in a stressful situations** | **0** | **12** | **5.73** | **+/-2.656** |
| **Q4- Score** | **The priority is the patient care** | **0** | **12** | **7.26** | **+/-2.403** |
| **Q5- Score** | **Communication skills** | **0** | **12** | **3.74** | **+/-2.817** |
| **Q6- Score** | **Teamwork** | **0** | **12** | **6.62** | **+/-3.094** |
| **Q7- Score** | **Ensuring patient safety** | **0** | **8** | **4.22** | **+/-2.452** |
| **Q8- Score** | **Work under pressure and ensure the safety of yourself and others** | **0** | **12** | **5.98** | **+/-2.621** |
| **Q9- Score** | **Respecting patient confidentiality** | **0** | **12** | **6.34** | **+/-2.97** |
| **Q10- Score** | **Ensuring patient safety** | **0** | **12** | **6.05** | **+/-2.717** |
| **TOTAL Score** |  | **28** | **88** | **55.52** | **+/-11.298** |

Moving on to the comparison of mean item scores between males and females (Table 4), it was observed that significant gender-based differences exist in participants’ responses to certain items. Notably, females tended to score in “Conflicts between professional and personal concerns” higher than males, with a mean score of 4.98 compared to 3.69,.This gender-based discrepancy was statistically significant (p = 0.006).both genders scored low regarding “communication skills” males scored 3.56 and females scored 3.93 and “ensuring patient safety” males scored 3.94 and females scored 4.52.However, for other items, no statistically significant differences were found.

**Table 4:**
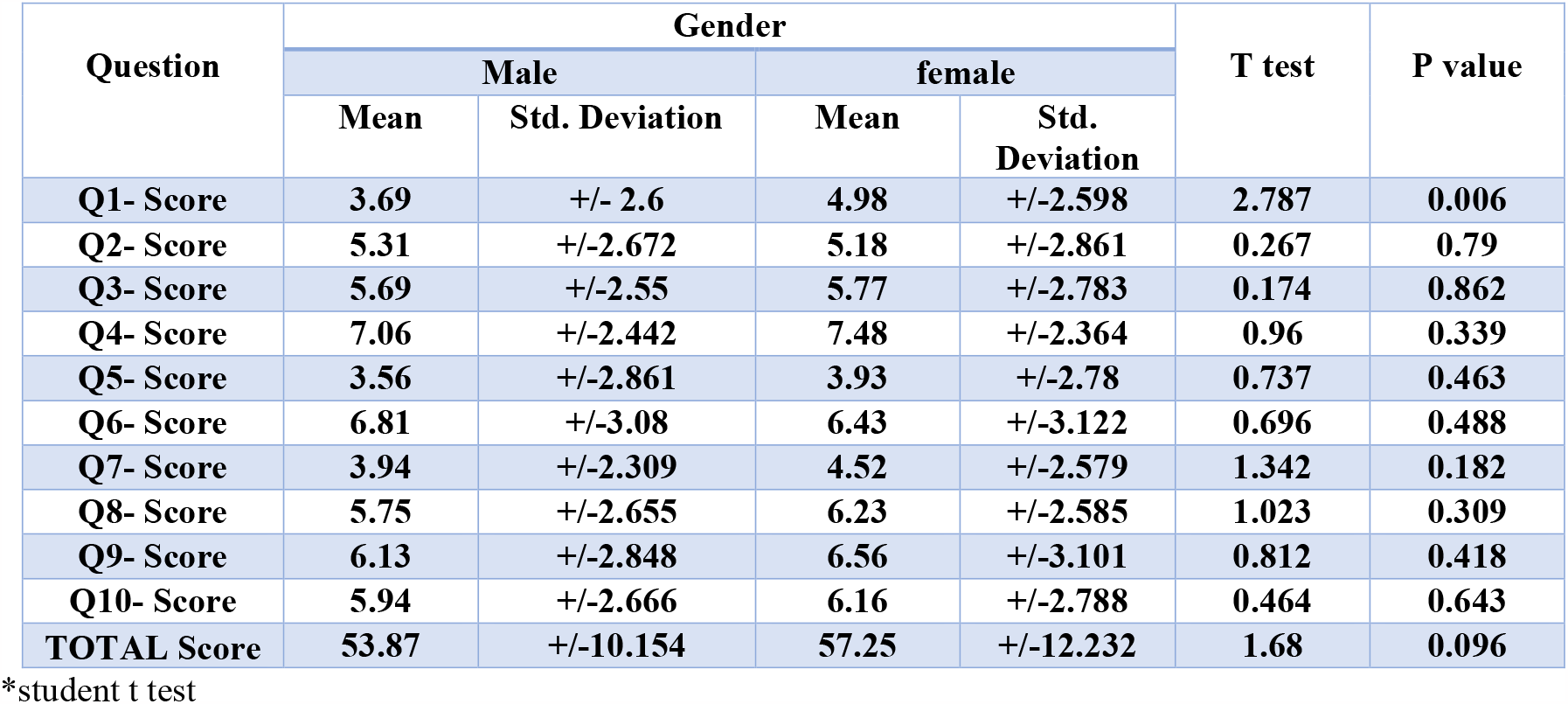
comparison of mean item score between males and females.

Furthermore, when assessing the impact of training level on participants’ responses (Table 5), the study revealed that rotators and senior residents differed significantly in their scores of several items. For instance, rotators tended to score higher on “Making decisions in stressful situations” and “ensuring patient safety “and “Work under pressure and ensure the safety of yourself and others,” while senior residents scored higher on “prescription error” and “Respecting patient confidentiality.” These differences were reflected in the total scores, with rotators having a mean total score of 57.6 compared to 53.6 for senior residents (p = 0.048).

**Table 5:** comparison of mean item score according to grade of training.

| Question | Training level |  |  |  | T test | P value |
| --- | --- | --- | --- | --- | --- | --- |
|  | Rotator |  | Senior resident |  |  |  |
|  | Mean | Std. Deviation | Mean | Std. Deviation |  |  |
| Q1- Score | 5.13 | +/-2.665 | 3.57 | +/-2.462 | 3.411 | 0.001 |
| Q2- Score | 5.07 | +/-2.742 | 5.42 | +/-2.778 | 0.706 | 0.482 |
| Q3- Score | 6.13 | +/-2.6 | 5.35 | +/-2.672 | 1.651 | 0.101 |
| Q4- Score | 7.27 | +/-2.269 | 7.26 | +/-2.539 | 0.012 | 0.991 |
| Q5- Score | 3.67 | +/-2.582 | 3.82 | +/-3.036 | 0.294 | 0.769 |
| Q6- Score | 6.73 | +/-2.991 | 6.52 | +/-3.207 | 0.378 | 0.706 |
| Q7- Score | 4.8 | +/-2.42 | 3.69 | +/-2.378 | 2.58 | 0.011 |
| Q8- Score | 6.33 | +/-2.582 | 5.66 | +/-2.636 | 1.438 | 0.153 |
| Q9- Score | 6.33 | +/-3.235 | 6.34 | +/-2.729 | 0.01 | 0.992 |
| Q10- Score | 6.13 | +/-2.896 | 5.97 | +/-2.562 | 0.336 | 0.737 |
| TOTAL Score | 57.6 | +/-11.719 | 53.6 | +/-10.625 | 2.001 | 0.048 |

\*student t test

In Table 6, a quartile-based analysis of gender and grade of training distribution of scores further highlighted the variation in scores among participants .The cut score between first quartile and second quartile was considered to represent the pass mark (48/120). The quartile analysis showed that female gender was associated with pass score compared to males had but the association was not statistically significant. It also showed that being senior resident was associated with pass score compared to rotators but the association was not statistically significant.

**Table 6:** Gender and grade of training distribution according to quartiles.

| Variables |  | Total Score |  | P value |
| --- | --- | --- | --- | --- |
|  |  | 48 or less<br>n =39 | more than 48<br>n =86 |  |
| Gender | Male<br>n= 64 | 22 | 42 | 0.4 |
|  | Female<br>n= 61 | 17 | 44 |  |
| Training Level | Rotator Resident<br>n= 60 | 18 | 42 | 0.7 |
|  | Senior Resident<br>n= 65 | 21 | 44 |  |

Table 7 provided insight into the comparison between training levels of the same gender in the total score. The data showed no significant differences in total scores between the training levels. This suggests that, despite variations in the scores on individual items, the overall assessment of their professional capabilities and challenges did not differ substantially between rotators and senior residents of the same gender.

**Table 7:** comparison between similar gender groups in total score.

| Gender | Training Level |  | t test | p value |
| --- | --- | --- | --- | --- |
|  | Rotator Resident | Senior Resident |  |  |
| Male | 55.64+/-10.8 | 52.9+/-9.75 | 0.98 | 0.32 |
| Female | 58.74+/-12.1 | 54.78+/-12.2 | 0.66 | 0.51 |

\*student t test

Lastly, Table 8 provides a comprehensive comparison of mean scores between males and females healthcare across the 2 groups, specifically categorized as “Rotator” and “Senior.” Within the Rotator category, the mean score for males is 55.64+/-10.8, while for females, it is 58.74+/-12.16. The student t-test statistics p-value of 0.32. This indicates that there is no statistically significant difference in mean scores between genders within the Rotator education level. Similarly, in the senior category, the mean score for males is 52.95+/-9.75, and for females, it is 54.78+/-12.2. The t-test statistics resulting in a p-value of 0.51. Consequently, there is no significant disparity in mean scores between males and females.

**Table 8:**
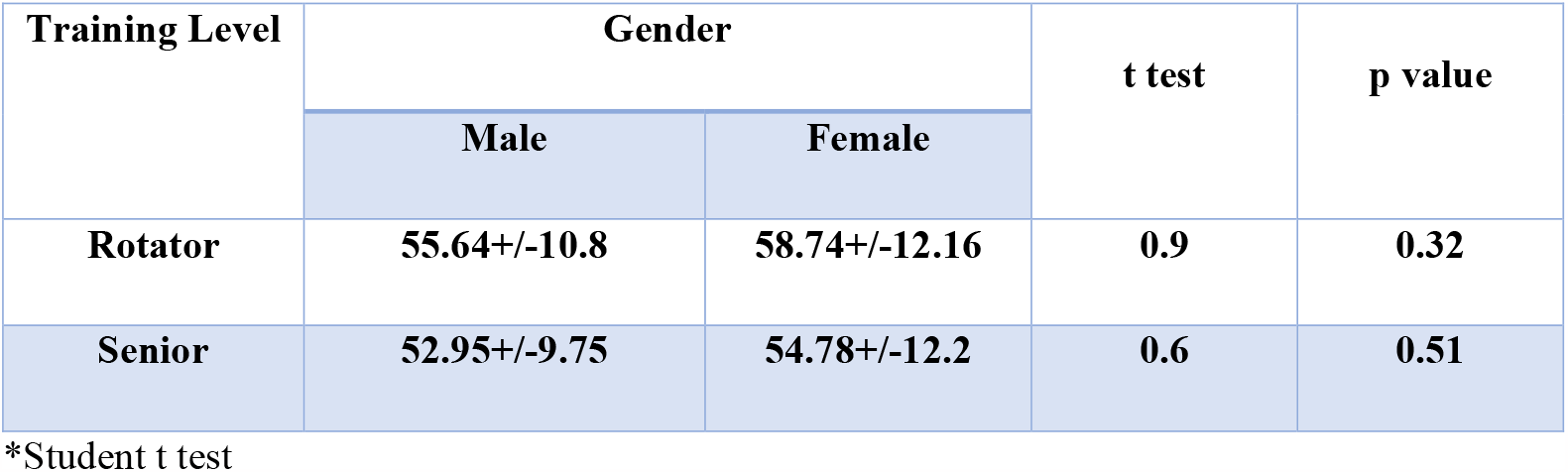
comparison between gender and education in total score.

## Discussion

A sample of 125 doctors participated in this study, in which 51% are male while 49% are female, and 48% of them are rotators and 52% are senior residents.

The data revealed variations in the mean scores across different aspects of their profession. Notably, “The priority is patient care” had the highest mean score of 7.26, indicating that participants highly prioritize patient care in their professional roles. On the other hand, “Communication skills” had a lower mean score of 3.74, suggesting a potential, although challenging, area for improvement in their abilities.

Barriers to effective communication include lack of confidence, lack of experience, complexity of healthcare, the distracting nature of healthcare settings, and lack of structure and standardization (28)(29)(30)(31)(32). The link between miscommunication and poor patient outcomes has been well documented (33).

Ineffective communication in healthcare results in delayed treatment, misdiagnosis, medication errors, patient injury, or death.

Improving the effectiveness of communication in healthcare is a global priority.

Interprofessional communication happens in synchronous and asynchronous means. Synchronous genres refer to communications happening in real time such as a meeting, ward round, handoff, or impromptu conversation (34). Communications also happen asynchronously such as on white boards, through medication orders, or written progress notes (34).

Communication is not only verbal and written, it includes body language, attitude and tone (30).

The literature suggests that physicians and nurses are trained differently in terms of communication styles and these differences lead to frustrations. Nurses are trained to be highly descriptive and physicians are trained to be succinct (35).

Physicians have noted frustration with nurse communications for “disorganization of information, illogical flow of content, lack of preparation to answer questions, inclusion of extraneous or irrelevant information, and delay in getting to the point” (36).

Nurses indicated concerns with physician communications due to “perceived inattentiveness especially during night hours, unwillingness to discuss goals of care, and feeling that a list of signs and symptoms had to be provided instead of just stating what the nurse thought the clinical problem was” (36).

There are various approaches for teaching interprofessional communication including workshops, online modules, and case studies. It is argued that sharing a common clinical experience such as simulation is a more effective approach compared to sitting together side-by-side in lecture halls (37).

Notably, females score in “Conflicts Between professional and personal concerns” higher than males, showing that Female were more careful about balancing professional and personal aspect and this was similar to another study conducted in India which found that majority of the women balance professional, personal, and social responsibilities equally (38).

Mean score of the question regarding of making decisions in stressful situations was slightly higher in female and this was different from another study conducted in Jamaica which found that females tend to make less utilitarian personal moral decisions compared to males, providing further evidence that there are gender differences in moral reasoning (39).

The priority is the patient care and Prescription error were closely similar in males and females similar to another study conducted in Saudi Arabia which found that the best score was given for reducing medical errors (6.2 points), followed by role of training and learning on patient safety (6 and 5.9 points). However, participants were not satisfied with undergraduate training on patient safety (4.8 points) (40).

Communication skills question score was low in both genders with females slightly higher than males while another study found that communication skills of males and females students were not significantly different (41).

The results revealed that mean score of males regarding teamwork was higher among them than female while another study conducted in Ecuador found that score was higher among female (42) Score of work under pressure and ensure the safety of yourself and others was higher among females than males and this was similar to another study found that score of females was higher (43). Respecting patient confidentiality score was higher among females than males, another study conducted in Spain found that both the female and male students showed high confidence as the clinical practicum progressed, facing these with greater ease and feeling more satisfied if the patients progressed well (44)

In Australia, a study found that unlike the medical student or the more senior doctor, the doctor in his or her early postgraduate years is simultaneously a responsible health professional, a subjugate learner and a human resource. These multiple roles generate the set of ethical issues faced by junior doctors, a set that has some overlaps with that faced by medical students and with that faced by more experienced doctors but is far from completely continuous with either (45).

Mean Score of prescription errors among senior residents was more than rotators, while another study conducted in UK found that calculated error rate of 38 errors made by junior staff per 1000 items prescribed by all doctors while residents had a reported error rate of 2 per 1000 items prescribed by all doctors (46). Mean Score for making decision among rotators was higher than that of senior resident, another study conducted in UK found that major stressors were effects of work on personal life, overwork, making mistakes, and making decisions especially among anesthetists (47). This finding may be related to the specialty of individuals and stressful events.

Both mean of communication skills and team work were higher among rotators than senior residents, while another study found that have also reported that inadequate communication skills and lack of teamwork of doctors were major obstacles to the provision of optimal patient care (48)

Rotators were more careful for patients’ safety than senior residents while another study conducted in USA found that senior doctors were more careful than junior (48).

Total score of male and female rotators was more than that of senior doctors and this was similar to another study conducted in Australia (49).This has several reasons, for example Work–life interference (or lack of balance), defined as an inter-role conflict where work demands make it such that one is unable to concurrently meet personal life demands or vice versa. The more individuals experience job demands, such as work overload and time pressure, the more work–life conflict they experience. While the direction of the conflict between work and life is bidirectional, the work and personal/family boundaries are easily permeable meaning that work demands tend to interfere with personal/family life to a greater extent than if the case was in reverse.

Variation in scores among participants, reflected the impact of their educational and gender-related experiences on their scores of the Profession. Among 39 participants with Total score <=48, 22 of them were male 17 were female (18 of them were Rotator and 21 were senior while 86 participants with Total score >48, 42 male residents and 44 senior female.

Regarding The comparison between gender and education in the total score, the data showed no significant differences in total scores between genders or training levels. This suggests that, despite variations in the scores on individual items, the overall assessment of their professional capabilities and challenges did not differ substantially bet between rotators and senior residents

There is no statistically significant difference in mean scores between genders within the Rotator training level. Similarly, in the senior category there is no significant disparity in mean scores between male and female healthcare professionals in the senior training level.

## CONCLUSIONS

This study tried to explore behavior and attitude toward complex and real time situations in work place. Mean score of conflicts between professional and personal concerns was higher for females. On the other hand regarding training level, there were two items significantly different between the 2 groups, namely conflicts between personal and professional concerns and ensuring patient safety. Communication skills had low score for both genders and training levels.

## Data Availability

All data produced in the present study are available upon reasonable request to the authors

## Recommendations

1. Further studies are recommended, that should include larger samples.
2. Increasing interest in the non-academic aspect of skills, specially communication skills.

## Limitations

Lack of cooperation from some of the senior residents and rotator residents.

